# Burden and Spectrum of Congenital Heart Disease in Nepal: A Systematic Review and Meta-Analysis

**DOI:** 10.1101/2025.09.20.25336248

**Authors:** Sandip Pandey, Apil Tiwari, Alisha Bhattarai, Anu Timalsina, Asmit Pandey, Aashis Poudel, Aakash Neupane, Deepak Jung Subedi

## Abstract

**Background:** Congenital heart disease (CHD) is the most common birth defect worldwide. In Nepal, reported prevalence ranges from less than 1 to over 100 per 1,000 live births, likely reflecting differences in study design rather than true variation. A synthesis accounting for study setting and size is needed to guide pediatric cardiac service planning.

**Methods:** We conducted a systematic review and meta-analysis following PRISMA guidelines (PROSPERO CRD420251125950). We searched PubMed, Embase, ScienceDirect, and Nepal-specific journals up to May 2025. Studies were included if they reported a CHD numerator and denominator from community, neonatal, or hospital settings. Prevalence was pooled using a binomial generalized linear mixed model. Pre-specified subgroups included study setting and geography. Atrial septal defect and ventricular septal defect proportions were pooled from patient-level data.

**Results:** Nine studies (N = 79,595; 462 CHD cases) were included. Overall pooled prevalence was 8.90 per 1,000 (95% CI 3.0 to 26.1; I² = 99.2%). Community studies yielded 3.81 per 1,000, and hospital studies yielded 25.64 per 1,000. After adjusting for sample size, the hospital effect was no longer significant. Larger studies consistently reported lower prevalence. ASD was the dominant lesion in community settings (53.7%), and VSD was more common in hospital settings (37.5%).

**Conclusions:** CHD prevalence in Nepal is highly context dependent. Community-based estimates are the most appropriate basis for health planning, while hospital figures reflect referral patterns rather than population burden. Nepal needs standardized population-based surveillance, broader echocardiography access outside Kathmandu, and clear referral pathways to address unmet surgical need.

**What Is Already Known About This Subject:** 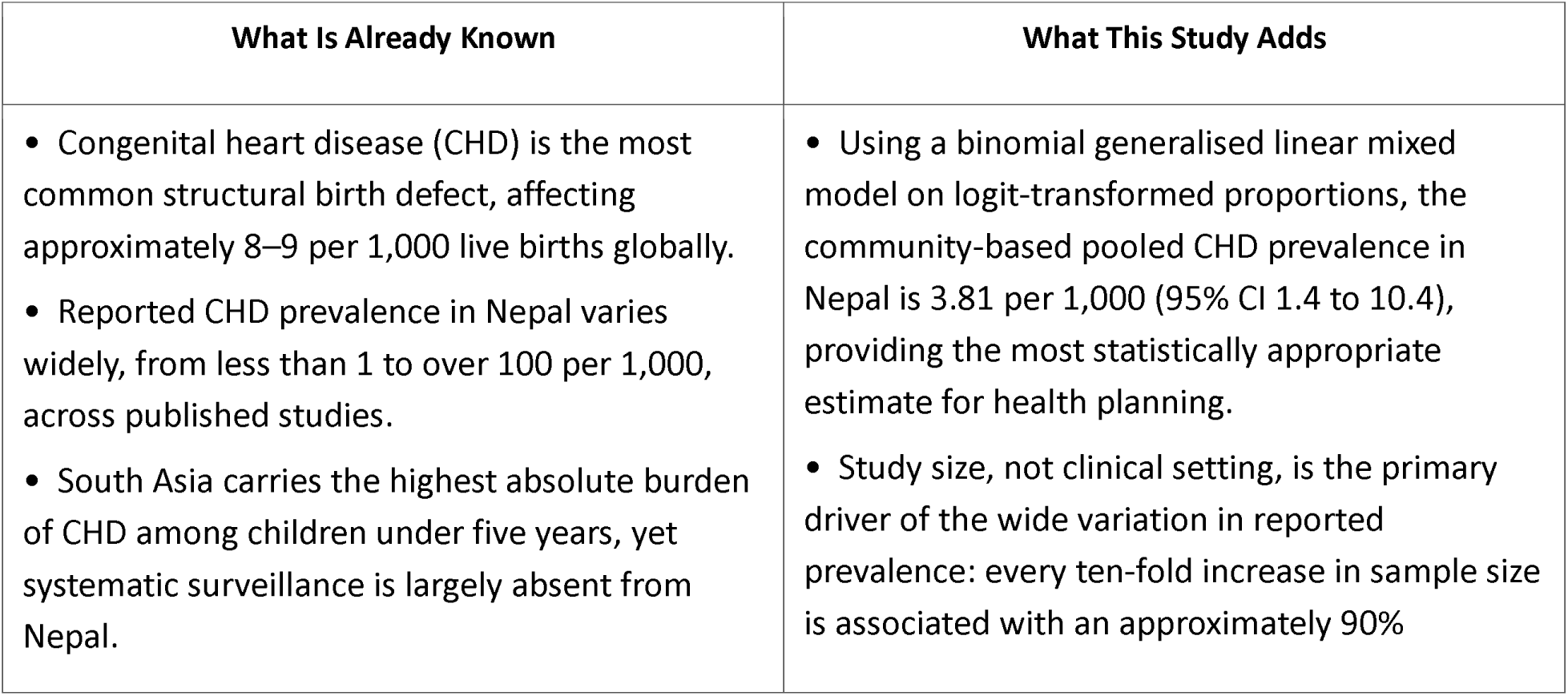

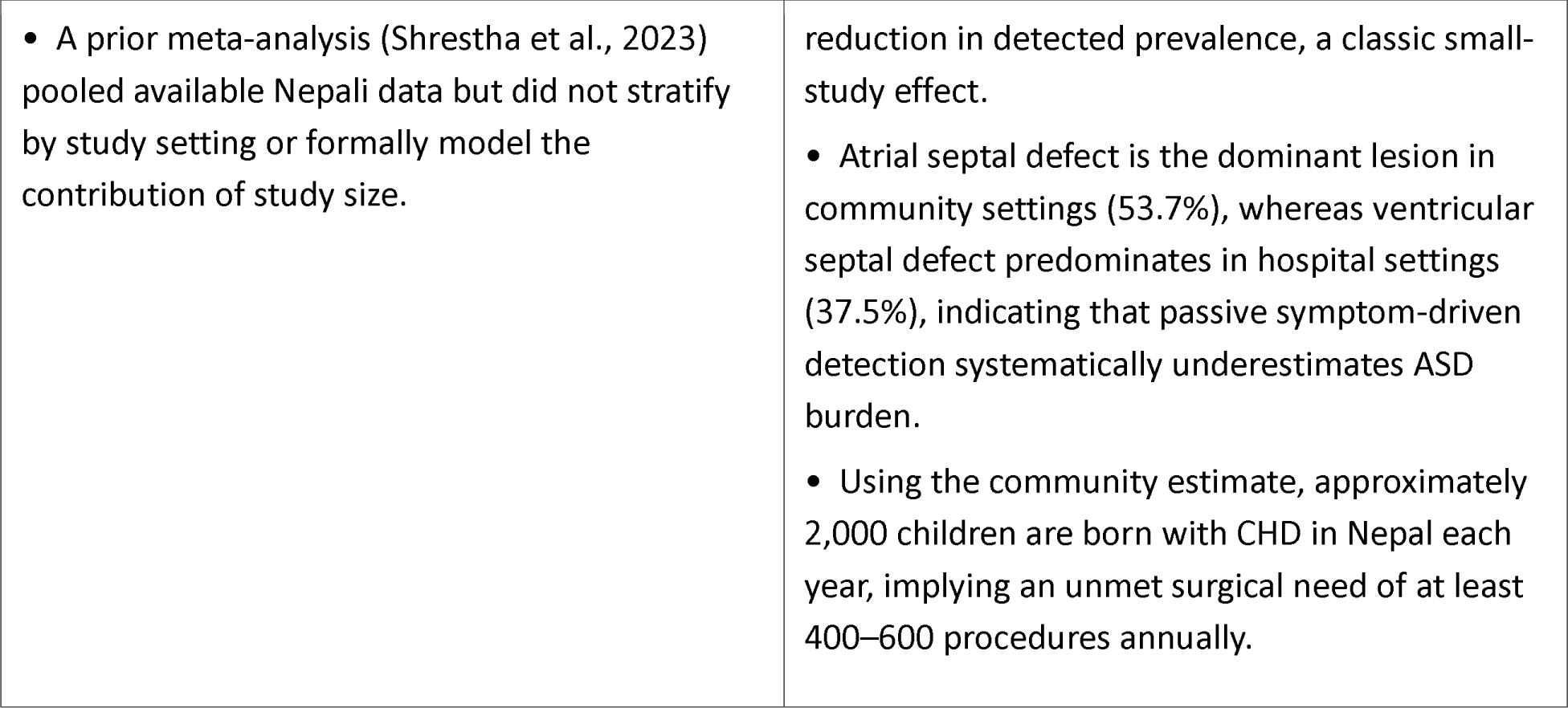

## Background

Congenital heart disease (CHD) refers to structural abnormalities of the heart or great vessels that are present at birth. These defects arise from incomplete or abnormal development of the fetal heart during early pregnancy. CHD encompasses a wide spectrum of conditions, ranging from simple, asymptomatic lesions that may resolve spontaneously to complex, life-threatening malformations requiring immediate medical or surgical intervention(1). The severity and clinical presentation of CHD vary significantly depending on the specific defect, its hemodynamic impact, and the presence of associated anomalies. Early diagnosis and appropriate management are crucial for improving outcomes and reducing morbidity and mortality associated with these conditions(2).

Globally, CHD represents a substantial public health challenge, affecting approximately 8 per 1,000 live births. The global birth prevalence of CHD has shown a progressive increase over recent decades, reaching an estimated 9.41 per 1,000 live births during 2010-2017 (3). While advancements in medical and surgical care have significantly improved survival rates in high-income countries, the burden of CHD remains disproportionately high in low- and middle-income countries (LMICs). These regions often face significant challenges, including limited access to diagnostic facilities, specialized healthcare professionals, and advanced treatment options, leading to higher rates of complications and mortality(4).

Within South Asia, the epidemiological landscape of CHD presents unique complexities. South Asia had the highest number of CHD cases globally, with India leading in absolute numbers(5). The prevalence of CHD in north-central India was found to be 19.14 per 1,000 individuals, with ventricular septal defect being the most common defect(6).Similarly, a retrospective analysis in the UK reported a higher estimated prevalence of CHD requiring hospital admission in Asian infants compared to non-Asian infants(7). Global data from 1990 to 2021 revealed that South Asia had the highest number of CHD cases among children under five years old(5). The challenges in this region are compounded by socioeconomic disparities, inadequate healthcare infrastructure, and a lack of comprehensive screening programs. These factors contribute to delayed diagnosis and suboptimal management, further exacerbating the impact of CHD on affected individuals and healthcare systems.

In Nepal, the true epidemiological burden of CHD remains largely undefined due to various systemic limitations. A significant portion of the population lives below the poverty line, and access to essential diagnostic services, such as echocardiography, is severely limited, particularly in rural areas. Existing data on CHD prevalence in Nepal are fragmented and often derived from single-center studies or limited regional surveys. For example, a systematic review and meta-analysis reported the prevalence of CHD in Nepal to be 0.7%, while other reports suggest a prevalence of 10.2% births(8) (9). These discrepancies highlight the urgent need for a more comprehensive and updated assessment.

This updated meta-analysis addresses these gaps in four ways. First, we synthesise pooled CHD prevalence across all eligible Nepal studies using a rigorous statistical model. Second, we quantify how much of the between-study variation is explained by study setting and study size. Third, we characterise the lesion spectrum (ASD and VSD) by setting. Fourth, we translate findings into specific implications for paediatric cardiac service planning in Nepal. Understanding the true community prevalence is a prerequisite for calculating unmet surgical need and for making the case for expanded paediatric cardiac infrastructure.

## Methods

### Protocol and registration

The protocol was registered with PROSPERO (CRD420251125950). Two amendments were made after registration: the search end date was extended to 1 May 2025, and the risk-of-bias tool was changed from the Newcastle-Ottawa Scale to the Joanna Briggs Institute (JBI) Prevalence Checklist. All other procedures followed the registered protocol. This report follows the PRISMA 2020 checklist.

### Search strategy and study selection

We searched PubMed, Embase, ScienceDirect, and Nepal-specific journals (Journal of Nepal Medical Association, Nepalese Heart Journal) from inception to 1 May 2025. The search string combined terms for CHD and epidemiology with a Nepal filter:

(“congenital heart disease” OR CHD OR “cardiac anomalies” OR “birth defects”) AND (prevalence OR epidemiology) AND (Nepal) NOT (“case reports”[Publication Type] OR “case series”[Title/Abstract] OR “meta-analysis”[Publication Type] OR “systematic review”[Publication Type])

There were no language restrictions. Results were exported to Rayyan for de-duplication. Two reviewers screened titles and abstracts independently. Disagreements were resolved by a third reviewer. Full texts were assessed against pre-specified eligibility criteria.

### Eligibility criteria

We included observational studies that reported CHD prevalence with an extractable numerator (CHD cases) and denominator (sample size) within one of three defined frames: community or school screenings, neonatal cohorts (live births or consecutive admissions), or hospital or referral cohorts including consecutive echocardiography series. We excluded rheumatic heart disease-only reports, case reports or series without denominators, convenience samples restricted to suspected CHD, studies without a clearly enumerated denominator, non-Nepal data, and duplicate or overlapping publications.

### Data extraction and quality assessment

Two reviewers independently extracted: first author, year, setting and region, sampling frame, age range, sample size (n), CHD cases, and lesion-level proportions for ASD and VSD. Setting was coded as community (CO) or hospital/referral (H). Geography was coded as Kathmandu Valley (urban) or sub-urban/mixed. Where only lesion percentages were reported, counts were reconstructed as round(% x CHD). Studies reporting multiple lesions per patient (percentages summing to more than 100%) were retained for overall CHD prevalence pooling but excluded from patient-level lesion pooling.

Two reviewers independently applied the JBI Critical Appraisal Checklist for Prevalence Studies across eight domains. Disagreements were resolved by discussion. Studies were classified qualitatively as low (seven or more “Yes” responses), moderate (four to six), or high (three or fewer) risk of bias.

### Statistical analysis

Prevalence was analysed on the logit scale. The primary model was a binomial GLMM (R package meta, sm = “PLOGIT”, random effects, method.tau = “ML”). Zero-cell handling used the binomial likelihood without continuity corrections; a small addition of 0.5 was applied only for degenerate strata. Where GLMM was unsupported for a subgroup, we used inverse-variance PLOGIT with restricted maximum likelihood (REML) and Hartung–Knapp (HK) confidence intervals as a fallback. Estimates were back-transformed and expressed per 1,000 persons. We report 95% CIs, 95% prediction intervals (PIs), and heterogeneity statistics (I² and τ² on the logit scale).

Pre-specified subgroups were study setting (CO versus H) and geography (Kathmandu Valley versus sub-urban/mixed). Lesion outcomes were synthesised as (1) share among CHD (ASD or VSD cases divided by total CHD cases) and (2) population prevalence (ASD or VSD cases divided by n), both using patient-level data only.

We performed exploratory random-effects meta-regression (R package metafor, Knapp–Hartung adjustment) with log₁₀(sample size) as the primary moderator and study setting as a secondary moderator. Publication year was examined in sensitivity models. Given k = 9, all meta-regression findings are hypothesis-generating. We performed leave-one-out sensitivity analyses and generated Baujat plots. Funnel plots and Egger tests were not performed because k was too small to yield reliable results; small-study effects were instead assessed via the log₁₀(n) moderator. GRADE certainty was assessed for all primary and lesion-level outcomes, accounting for risk of bias, inconsistency, imprecision, indirectness, and publication bias.

## Results

### Study selection

We identified 27 records (25 from databases, 2 from other sources). After removing 8 duplicates, we screened 17 titles and abstracts and excluded 8. We assessed 9 full texts for eligibility. Two studies Gautam NC (2021)(10) and Joshi A(2016)(11) were excluded because they were descriptive reports without extractable denominators. Nine studies met all inclusion criteria (N = 79,595; CHD cases = 462). The PRISMA flow diagram is shown in Figure 1.

**Figure 1.** PRISMA 2020 flow diagram. Schematic overview of study identification, screening, eligibility assessment, and inclusion. Records were identified from PubMed, Embase, ScienceDirect, and Nepal-specific journals (n = 25) and from other sources (n = 2). After removal of 8 duplicates, 17 titles and abstracts were screened. Eight records were excluded at title/abstract stage. Nine full texts were assessed for eligibility. Two were excluded because they were descriptive reports without extractable denominators (Gautam 2021, Joshi 2016). Nine studies were included in the final meta-analysis. CHD = congenital heart disease; PRISMA = Preferred Reporting Items for Systematic Reviews and Meta-Analyses.

### Study characteristics

Table 1 summarises the characteristics of the nine included studies. They span four decades (2003 to 2024) and cover Kathmandu Valley, Sunsari, Kavre, and Dharan. Five studies used community or school frames (CO) and four used hospital or referral frames (H). Sample sizes ranged from 219 (Dongol SS, 2018) to 34,876 (Prajapati D, 2013). Diagnostic methods varied from clinical auscultation with selective echocardiography to consecutive echocardiography for all participants. Two studies (Chapagain RH, 2017; Dongol SS, 2018) reported lesion proportions summing to more than 100% and were excluded from patient-level lesion pooling.

**Table 1.** Characteristics of the nine included studies.

| First Author | Year of Publication | Region/Province | Sample Size (n) | Number of CHD Cases | Prevalence per 1,000 | ASD (%) | VSD (%) | Setting |
| --- | --- | --- | --- | --- | --- | --- | --- | --- |
| Chapagain RH(9) | 2017 | Kathmandu | 831 | 85 | 102.28 | 87.1<sup>†</sup> | 16.5<sup>†</sup> | H |
| Prajapati D(12) | 2013 | Kathmandu | 34876 | 38 | 1.09 | 47.4 | 28.9 | CO |
| Dongol SS(13) | 2018 | Kavre | 219 | 29 | 132.00 | 6.8 | 31 | H |
| KC Man Bahadur(14) | 2003 | Kathmandu | 9420 | 12 | 1.30 | 50 | 25 | CO |
| Koirala PC(15) | 2024 | Kathmandu | 3593 | 30 | 8.30 | 63.33 | 13.33 | CO |
| Shrestha S(16) | 2020 | Kathmandu | 2456 | 12 | 4.90 | 66.7 | 16.7 | H |
| Shah GS(17) | 2008 | Dharan | 14461 | 84 | 5.80 | 4.8 | 58.3 | H |
| Shakya S(18) | 2011 | Multiple | 7166 | 150 | 20.90 | na | na | CO |
| Shrestha N(19) | 2021 | Sunsari | 6573 | 22 | 3.30 | na | na | CO |

<sup>†</sup> Multi-lesion reporting: percentages are expressed as 'proportion of lesions' and may sum to more than 100%; these studies were excluded from patient-level lesion pooling. ASD = atrial septal defect; VSD = ventricular septal defect; CHD = congenital heart disease; CO = community/school setting; H = hospital/referral setting; NA = not available.

### Risk of bias

Table 6 presents domain-level JBI judgements. Community studies had some concern in most domains, driven by mixed randomisation procedures and incomplete response rate reporting. Hospital studies had serious concern for sampling frame and coverage, because hospital-based recruitment does not represent the community population. No study was judged at low risk of bias overall. The body of evidence carries moderate to high risk of bias.

### Overall CHD prevalence

The overall pooled CHD prevalence was 8.90 per 1,000 (95% CI 3.0 to 26.1; 95% PI 0.2 to 338.9; I^2^ = 99.2%, tau^2^ = 2.765 on the logit scale). Heterogeneity was extreme. The wide prediction interval indicates that in a new Nepali study drawn from the same evidence base, the true prevalence could plausibly range from near zero to more than 300 per 1,000, depending on the sampling frame and study size. The forest plot is presented as Figure 2 and subgroup results are summarised in Table 2.

**Figure 2.** Forest plot of overall CHD prevalence (per 1,000, logit scale, zoomed axis). Random-effects binomial GLMM model. Each row shows the study name, total sample size (N), CHD cases, and the back-transformed proportion with 95% CI. The diamond represents the pooled estimate (8.90 per 1,000; 95% CI 3.0 to 26.1). The horizontal bar below the diamond represents the 95% PI (0.2 to 338.9). Heterogeneity: I² = 99.2%, τ² = 2.765 (logit scale). CHD = congenital heart disease; CI = confidence interval; CO = community/school setting; GLMM = generalised linear mixed model; H = hospital/referral setting; I² = Higgins heterogeneity statistic; PI = prediction interval; τ² = between-study variance on the logit scale.

### Subgroup analyses

#### By study setting

Community studies (k = 5) yielded a pooled prevalence of 3.81 per 1,000 (95% CI 1.4 to 10.4; I² = 98.8%). Hospital/referral studies (k = 4) yielded 25.64 per 1,000 (95% CI 5.4 to 113.2; I² = 99.4%). The approximately seven-fold descriptive difference reflects referral bias and more intensive diagnostic ascertainment in clinical settings, not a true difference in disease frequency. After adjusting for log₁₀(sample size) in meta-regression, the hospital versus community effect attenuated substantially and was no longer statistically significant (odds ratio [OR] 1.42; 95% CI not reliably estimable; see Table 5). Forest plots for community and hospital subgroups are presented as Figures 3a and 3b.

**Figure 3a.** Forest plot of CHD prevalence in community studies (per 1,000, zoomed axis). Five community/school studies. Pooled estimate: 3.81 per 1,000 (95% CI 1.4 to 10.4; 95% PI 0.1 to 107.0). I² = 98.8%, τ² = 1.275. Model: binomial GLMM, logit scale. CHD = congenital heart disease; CI = confidence interval; GLMM = generalised linear mixed model; I² = Higgins heterogeneity statistic; PI = prediction interval; τ² = between-study variance.

**Figure 3b.** Forest plot of CHD prevalence in hospital/referral studies (per 1,000, zoomed axis). Four hospital/referral studies. Pooled estimate: 25.64 per 1,000 (95% CI 5.4 to 113.2; 95% PI 0.1 to 887.1). I² = 99.4%, τ² = 2.558. Model: binomial GLMM, logit scale. CHD = congenital heart disease; CI = confidence interval; GLMM = generalised linear mixed model; I² = Higgins heterogeneity statistic; PI = prediction interval; τ² = between-study variance.

**Table 2.** Pooled CHD prevalence by subgroup.

| Subgroup | k | Pooled per 1,000 | 95% CI | 95% PI | $I^2$ (%) | $\tau^2$ |
| --- | --- | --- | --- | --- | --- | --- |
| Overall (all studies) | 9 | 8.90 | 3.0 to 26.1 | 0.2 to 338.9 | 99.2 | 2.765 |
| Community (CO) | 5 | 3.81 | 1.4 to 10.4 | 0.1 to 107.0 | 98.8 | 1.275 |
| Hospital/referral (H) | 4 | 25.64 | 5.4 to 113.2 | 0.1 to 887.1 | 99.4 | 2.558 |
| Kathmandu Valley (urban) | 5 | 5.69 | 1.3 to 24.8 | 0.03 to 493.2 | 99.4 | 2.845 |
| Sub-urban/mixed | 4 | 15.46 | 3.7 to 61.7 | 0.1 to 734.8 | 98.9 | 2.108 |

Estimates are back-transformed from the logit scale. Primary model: binomial GLMM; fallback: inverse-variance PLOGIT with Hartung–Knapp correction. CHD = congenital heart disease; CI = confidence interval; CO = community/school setting; GLMM = generalised linear mixed model; H = hospital/referral setting; $I^2$ = Higgins heterogeneity statistic; k = number of studies; PI = prediction interval; PLOGIT = logit transformation for proportions; $\tau^2$ = between-study variance on the logit scale.

### By geography

Kathmandu Valley studies (k = 5) yielded a pooled prevalence of 5.69 per 1,000 (95% CI 1.3 to 24.8; I² = 99.4%). Sub-urban and mixed-region studies (k = 4) yielded 15.46 per 1,000 (95% CI 3.7 to 61.7; I² = 98.9%). The higher estimate in sub-urban settings partly reflects the inclusion of hospital-based studies with very small sample sizes (for example, Dongol SS, 2018: n = 219) rather than a genuine geographic excess. Forest plots are presented as Figures 4a and 4b.

**Figure 4a.** Forest plot of CHD prevalence in Kathmandu Valley studies (per 1,000, zoomed axis). Five Kathmandu Valley studies. Pooled estimate: 5.69 per 1,000 (95% CI 1.3 to 24.8; 95% PI 0.03 to 493.2). I² = 99.4%, τ² = 2.845. CHD = congenital heart disease; CI = confidence interval; I² = Higgins heterogeneity statistic; PI = prediction interval; τ² = between-study variance.

**Figure 4b.** Forest plot of CHD prevalence in sub-urban/mixed-region studies (per 1,000, zoomed axis). Four sub-urban or mixed-region studies. Pooled estimate: 15.46 per 1,000 (95% CI 3.7 to 61.7; 95% PI 0.1 to 734.8). I² = 98.9%, τ² = 2.108. CHD = congenital heart disease; CI = confidence interval; I² = Higgins heterogeneity statistic; PI = prediction interval; τ² = between- study variance.

### Lesion-level results

Table 3 presents ASD and VSD shares among CHD cases (patient-level data only; k = 3 per cell after excluding multi-lesion studies). In community settings, ASD was the dominant lesion (pooled share 53.7%, 95% CI 42.8% to 64.3%), while VSD accounted for 22.5% (95% CI 14.7% to 32.9%) with low heterogeneity. In hospital settings, VSD share increased to 37.5% (95% CI 19.1% to 60.5%) and ASD share fell to 15.2% (95% CI 2.4% to 56.5%), with high heterogeneity in both. This pattern is consistent with referral of haemodynamically significant, auscultation-positive lesions (predominantly large VSDs) to hospital, while small ASDs that are clinically silent are detected mainly through active community screening. Forest plots for VSD share in community and hospital settings are shown in Figures 5a and 5b.

Table 4 presents population-level lesion prevalence. Community estimates were 1.18 per 1,000 for ASD and 0.38 per 1,000 for VSD. Hospital estimates were 4.93 per 1,000 for ASD and 6.68 per 1,000 for VSD. All lesion-level analyses were associated with wide prediction intervals and very low to low certainty (Table 7).

**Figure 5a.** Forest plot of VSD share among CHD cases in community studies (proportion, zoomed axis). Three patient-level community studies. Pooled VSD share: 22.5% (95% CI 14.7% to 32.9%; 95% PI 8.4% to 47.9%). I² = 12.9%, τ² = 0. Low heterogeneity. CHD = congenital heart disease; CI = confidence interval; CO = community/school setting; I² = Higgins heterogeneity statistic; PI = prediction interval; VSD = ventricular septal defect; τ² = between-study variance.

**Figure 5b.** Forest plot of VSD share among CHD cases in hospital studies (proportion, zoomed axis). Three patient-level hospital studies. Pooled VSD share: 37.5% (95% CI 19.1% to 60.5%; 95% PI 1.8% to 95.2%). I² = 81.1%, τ² = 0.434. Substantial heterogeneity. CHD = congenital heart disease; CI = confidence interval; H = hospital/referral setting; I² = Higgins heterogeneity statistic; PI = prediction interval; VSD = ventricular septal defect; τ² = between-study variance.

**Table 3.** ASD and VSD share among CHD cases by setting (patient-level studies only).

| Setting | Lesion | k | Pooled (%) | 95% CI (%) | 95% PI (%) | $I^2$ (%) | $\tau^2$ |
| --- | --- | --- | --- | --- | --- | --- | --- |
| Community (CO) | ASD | 3 | 53.7 | 42.8 to 64.3 | 30.7 to 75.3 | 0 | 0 |
| Community (CO) | VSD | 3 | 22.5 | 14.7 to 32.9 | 8.4 to 47.9 | 12.9 | 0 |
| Hospital/referral | ASD | 3 | 15.2 | 2.4 to 56.5 | 0.0 to 99.9 | 91.4 | 2.676 |

| Setting | Lesion | k | Pooled (%) | 95% CI (%) | 95% PI (%) | I<sup>2</sup> (%) | τ<sup>2</sup> |
| --- | --- | --- | --- | --- | --- | --- | --- |
| (H) |  |  |  |  |  |  |  |
| Hospital/referral (H) | VSD | 3 | 37.5 | 19.1 to 60.5 | 1.8 to 95.2 | 81.1 | 0.434 |

Multi-lesion studies (percentages summing to more than 100% of CHD cases) were excluded from patient-level pooling. ASD = atrial septal defect; CHD = congenital heart disease; CI = confidence interval; CO = community/school setting; H = hospital/referral setting; I<sup>2</sup> = Higgins heterogeneity statistic; k = number of studies; PI = prediction interval; VSD = ventricular septal defect; τ<sup>2</sup> = between-study variance on the logit scale.

**Table 4.** Population prevalence of ASD and VSD by setting (per 1,000; patient-level studies only).

| Setting | Lesion | k | Pooled per 1,000 | 95% CI | 95% PI | I<sup>2</sup> (%) | τ<sup>2</sup> |
| --- | --- | --- | --- | --- | --- | --- | --- |
| Community (CO) | ASD | 3 | 1.18 | 0.35 to 4.00 | 0.006 to 178.7 | 0 | 0 |
| Community (CO) | VSD | 3 | 0.38 | 0.24 to 0.60 | 0.14 to 1.04 | 12.9 | 0 |
| Hospital/referral (H) | ASD | 3 | 4.93 | 0.55 to 44.1 | 0.001 to >999 | 91.4 | 2.676 |
| Hospital/referral (H) | VSD | 3 | 6.68 | 1.58 to 28.3 | 0.04 to >999 | 81.1 | 0.434 |

Multi-lesion studies excluded from patient-level pooling. ASD = atrial septal defect; CI = confidence interval; CO = community/school setting; H = hospital/referral setting; I<sup>2</sup> = Higgins heterogeneity statistic; k = number of studies; PI = prediction interval; VSD = ventricular septal defect; τ<sup>2</sup> = between-study variance on the logit scale.

Table 5 presents meta-regression results. There was a strong, consistent inverse association between study size and reported prevalence: for every ten-fold increase in sample size, the odds of CHD detection fell by approximately 90% (OR approximately 0.10). This is a hallmark of small-study effects. Small, often hospital-based studies with intensive targeted echocardiography reported very high prevalence. Large, population-based surveys with less sensitive screening reported conservative estimates. The unadjusted hospital versus community OR of 6.89 attenuated to 1.42 after adjusting for log₁₀(n), and was not statistically significant, indicating that study size rather than clinical setting drives most of the raw difference. No secular trend with publication year was identified.

**Table 5.** Random-effects meta-regression results (Knapp–Hartung adjustment).

| Predictor | log-OR | Lower 95% CI | Upper 95% CI | OR | Model |
| --- | --- | --- | --- | --- | --- |
| $\log_{10}(\text{sample size})$ [primary] | -2.28 | Not estimable | Not estimable | 0.10 | $\log_{10}(n)$ only |
| Setting: hospital vs community [unadjusted] | 1.93 | Not estimable | Not estimable | 6.89 | Unadjusted |
| Setting: hospital vs community [adj. $\log_{10}(n)$] | 0.35 | Not estimable | Not estimable | 1.42 | Adjusted for n |
| $\log_{10}(\text{sample size})$ [adj. setting] | -2.12 | Not estimable | Not estimable | 0.12 | Adjusted for setting |

Given $k = 9$ overall ( $k = 3$ to $5$ per subgroup), confidence intervals were not reliably estimable for all individual terms. All meta-regression results are hypothesis-generating. CI = confidence interval; log-OR = log-transformed odds ratio; OR = odds ratio.

### Sensitivity Analyses

Leave-one-out analyses showed modest variation in pooled estimates across all strata. For all studies combined, τ² ranged from 2.01 to 3.11 and I² remained between 98.8% and 99.3% regardless of which study was omitted. No single study was decisively influential. The Baujat plot (Figure 6) showed diffuse contributions to heterogeneity. Results from GLMM and inverse- variance plus Hartung–Knapp were qualitatively identical. The leave-one-out forest plot is presented as Figure 7.

**Figure 6.** Baujat plot of contribution to heterogeneity versus influence on the overall pooled estimate. Each circle represents one study. The x-axis shows each study’s contribution to the overall Q statistic. The y-axis shows influence on the overall pooled result. No single study dominates either axis, indicating that heterogeneity is diffuse across the evidence base. CHD = congenital heart disease.

**Figure 7.** Leave-one-out forest plot for the overall CHD prevalence estimate. Each row shows the pooled estimate with 95% CI and 95% PI after omitting the labelled study. The diamond at the bottom shows the full model. Estimates and heterogeneity statistics are stable across omissions, confirming the robustness of conclusions. CHD = congenital heart disease; CI = confidence interval; PI = prediction interval.

### Risk of Bias Assessment

Body-of-evidence appraisal with the JBI prevalence checklist indicated moderate-to-high risk of bias, driven by non-population sampling frames and uneven coverage in hospital/referral cohorts, and by inconsistent ascertainment across designs.

**Table 6.** JBI Critical Appraisal domain-level judgements by setting. *JBI domain-level judgements here — two rows (CO studies; H studies) × eight JBI domains. Colour code: low concern = green; some concern = yellow; serious concern = red.* CO = community/school setting; H = hospital/referral setting. Judgements are qualitative (low concern, some concern, serious concern) based on JBI Prevalence Checklist domains. No study reached low risk of bias overall.

### Certainty of Evidence

Given extreme heterogeneity and imprecision, GRADE certainty for prevalence outcomes was judged very low, with lesion-share estimates ranging from low to very low depending on subgroup.

**Table 7.** GRADE Summary of Findings for key outcomes. *GRADE Summary of Findings. Columns: Outcome | k | Pooled estimate | 95% CI | 95% PI | Certainty | Reason for downgrading* All prevalence outcomes were downgraded for risk of bias (sampling frames), inconsistency (extreme heterogeneity), and imprecision (wide prediction intervals). Certainty ranges from low (VSD share, community setting) to very low (all other outcomes). CHD = congenital heart disease; CI = confidence interval; CO = community/school setting; GRADE = Grading of Recommendations, Assessment, Development and Evaluations; H = hospital/referral setting; k = number of studies; PI = prediction interval.

## Discussion

### Principal findings

This meta-analysis of nine Nepali studies shows that reported CHD prevalence is determined more by where cases are counted than by how common CHD actually is. The seven-fold gap between community (3.81 per 1,000) and hospital estimates (25.64 per 1,000) is the expected imprint of referral bias and intensive diagnostic ascertainment in clinical settings. After adjusting for study size, this gap largely disappears. Large community studies consistently report lower prevalence than small hospital series. This pattern is a classic signal of small-study effects: smaller studies, usually hospital-based with targeted echocardiography, detect more disease per person examined.

The community estimate of 3.81 per 1,000 falls within the global range for moderate-income South Asian populations. For comparison, community prevalence in north-central India was estimated at 19.14 per 1,000 (though that study used school-based echocardiography, which detects more lesions than clinical screening), and a Malaysian population-based study reported 6.7 per 1,000.6,7 The lower community estimate in Nepal most likely reflects lower echocardiographic coverage in screening studies rather than a genuinely lower burden.

The ASD-dominant community profile and VSD-dominant hospital profile are clinically informative. Large VSDs cause haemodynamically significant left-to-right shunts detectable on auscultation, so affected children are more likely to reach hospital. Small ASDs are often asymptomatic and silent on auscultation, so they are detected only during active echocardiographic screening. This pattern confirms that passive, symptom-driven detection systematically underestimates ASD burden in Nepal.

### Implications for paediatric cardiac services in Nepal

These findings carry direct implications for how Nepal plans and delivers cardiac care for children.

#### Estimating unmet surgical need

Using the community estimate of 3.81 per 1,000 and Nepal’s annual birth cohort of approximately 530,000 live births, approximately 2,000 children are born with CHD in Nepal each year. Of these, approximately 20% to 30% will require catheter-based or surgical intervention in infancy or early childhood. That translates to a minimum of 400 to 600 procedures needed per year in the paediatric age group alone. Current capacity at Shahid Gangalal National Heart Center and Manmohan Cardiothoracic Vascular and Transplant Center falls well short of this estimate. The unmet need is large and growing.

#### Neonatal and infant screening

The community ASD share of 53.7% confirms that a large proportion of CHD in Nepal is clinically silent in the newborn period. Pulse oximetry screening, which reliably detects cyanotic defects, will miss the majority of acyanotic lesions (ASD, small VSD). Nepal should consider adding echocardiographic evaluation to high-risk newborn assessments (low birth weight, trisomy, maternal diabetes, rubella) as a first step toward systematic screening, even before universal echocardiographic screening is feasible.

#### Expanding echocardiography outside Kathmandu

The higher pooled prevalence in sub-urban and mixed regions (15.46 per 1,000) partly reflects small-study bias. However, the near-absence of echocardiography in rural Nepal means that many affected children in these areas will never receive a diagnosis. Training programmes to upskill community health officers in focused cardiac ultrasound, supported by telemedicine interpretation from tertiary centres, represent a feasible intermediate step.

#### Referral pathway design

The ASD-VSD gradient by setting suggests that current referral pathways capture the symptomatic, larger lesions while missing the smaller, clinically silent ones. A structured referral protocol based on clinical examination by trained paediatric nurses, with clear thresholds for echocardiographic referral, could improve detection without requiring universal echocardiography at primary level.

### Comparison with prior meta-analyses

A prior systematic review by Shrestha et al. (2023) reported a pooled CHD prevalence of 0.7% in Nepal(8). The present study supersedes that analysis in several ways. First, we include two additional recent studies (Koirala 2024, Shrestha 2021). Second, we use a binomial GLMM on the logit scale rather than simple inverse-variance pooling, which handles extreme heterogeneity and proportion data more appropriately. Third, we separate community and hospital estimates and formally model the contribution of study size, revealing that small-study effects account for much of the wide range. Fourth, we provide 95% prediction intervals, which give a more honest representation of between-study variability than confidence intervals alone. The GRADE certainty ratings (very low for all prevalence outcomes) reflect the limitations of the primary evidence and should not be interpreted as a weakness of the analysis.

### Heterogeneity

Extreme heterogeneity (I2 = 99%) is not a flaw of this analysis. It is an honest description of the primary evidence base. When study frames range from 219-person neonatal cohorts with echocardiography of all admissions to 34,876-person school screenings with auscultation alone, wide variation is expected and informative. The meta-regression identifies study size as the main statistical driver of this variation. Residual heterogeneity after adjusting for size and setting likely reflects differences in echocardiographic protocols, operator skill, and lesion classification thresholds across studies. This residual variation is captured by the prediction intervals and should be communicated to policymakers as irreducible uncertainty in the current evidence base.

### Strengths and limitations

Strengths of this review include a comprehensive multi-database search, use of advanced statistical models (GLMM), pre-specified stratification by setting and geography, patient-level lesion synthesis with exclusion of multi-lesion-reporting studies, meta-regression that quantifies key drivers of heterogeneity, and multiple robustness checks. GRADE-based certainty assessment provides transparent communication of evidence quality.

Limitations are inherent to the primary evidence base. Only nine studies were eligible. Several subgroup analyses used k = 3 to 5 studies, making meta-regression severely underpowered and all subgroup conclusions provisional. Sampling frames were heterogeneous. Lesion counts were reconstructed from reported percentages in some studies. Echocardiographic protocols were not standardised across studies. Risk of bias was moderate to high in all studies. Funnel plot-based tests for publication bias could not be performed. The certainty of all prevalence estimates is very low. These limitations reinforce the urgent need for a prospective, standardised, population-based national CHD surveillance programme in Nepal.

### Conclusions

CHD prevalence in Nepal is not a single number. It is a range that depends critically on where cases are ascertained and how large the study is. Community-based estimates (3.81 per 1,000) are the most appropriate anchor for public health planning and suggest that approximately 2,000 children with CHD are born in Nepal each year. Hospital estimates reflect referral case-mix and clinical service demand, not population burden.

Nepal’s paediatric cardiac services face a large and growing unmet need. Addressing this requires three parallel investments: establishing a standardised, population-based CHD surveillance programme; expanding echocardiographic capacity and training outside Kathmandu Valley; and developing clear referral pathways from primary care to the two existing paediatric cardiac surgical centres. Without these steps, most Nepali children born with CHD will continue to go undiagnosed until they present with advanced disease.

## Data Availability

All data produced in the present study are available upon reasonable request to the authors

## Abbreviations

ASD: Atrial septal defect
CHD: Congenital heart disease
CI: Confidence interval
CO: Community study setting
GLMM: Generalised linear mixed model
GRADE: Grading of Recommendations, Assessment, Development and Evaluations
H: Hospital or referral setting
HK: Hartung–Knapp
I²: Higgins heterogeneity statistic
JBI: Joanna Briggs Institute
LMIC: Low- and middle-income country
ML: Maximum likelihood
n: Study sample size
N: Total pooled sample size
OR: Odds ratio
PI: Prediction interval
PLOGIT: Logit transformation for proportions
PRISMA: Preferred Reporting Items for Systematic Reviews and Meta-Analyses
PROSPERO: International Prospective Register of Systematic Reviews
REML: Restricted maximum likelihood
RHD: Rheumatic heart disease
RoB: Risk of bias
τ²: Between-study variance on the logit scale
VSD: Ventricular septal defect

## Declarations

### Ethical approval

Not applicable. This meta-analysis is based exclusively on previously published data and does not involve human or animal participation.

### Consent for publication

Not applicable. This manuscript does not contain individual participant data.

### Data availability

Derived tables, figures (CSV/Excel/PDF), and R analysis code are available on reasonable request from the corresponding author.

### Authors’ contributions (CRediT)

Conceptualisation, methodology, guarantor: Sandip Pandey. Screening, data curation: Apil Tiwari, Alisha Bhattarai. Risk of bias assessment: Anu Timalsina. Data extraction, figures: Asmit Pandey. Formal analysis, visualisation: Aakash Neupane. Supervision, methodology: Deepak Jung Subedi. Writing (original draft): Sandip Pandey. Writing (review and editing): all authors.

